# INDIA’S PRAGMATIC VACCINATION STRATEGY AGAINST COVID-19: A MATHEMATICAL MODELLING BASED ANALYSIS

**DOI:** 10.1101/2021.05.07.21256742

**Authors:** Sandip Mandal, Nimalan Arinaminpathy, Balram Bhargava, Samiran Panda

**Affiliations:** Indian Council of Medical Research, New Delhi, India; MRC Centre for Global Infectious Disease Analysis, School of Public Health, Imperial College London, London, UK

## Abstract

**Objectives:** To investigate the impact of targeted vaccination strategies on morbidity and mortality due to COVID-19, as well as on the incidence of SARS-CoV-2, in India.

**Design:** Mathematical modelling.

**Settings:** Indian epidemic of COVID-19 and vulnerable population.

**Data sources:** Country specific and age-segregated pattern of social contact, case fatality rate and demographic data obtained from peer-reviewed literature and public domain.

**Model:** An age-structured dynamical model describing SARS-CoV-2 transmission in India incorporating uncertainty in natural history parameters was constructed.

**Interventions:** Comparison of different vaccine strategies by targeting priority groups such as key workers including health care professionals, individuals with comorbidities (24 – 60 year), and all above 60.

**Main outcome measures:** Incidence reduction and averted deaths in different scenarios, assuming that the current restrictions are fully lifted as vaccination is implemented.

**Results:** The priority groups together account for about 18% of India’s population. An infection preventing vaccine with 60% efficacy covering all these groups would reduce peak symptomatic incidence by 20.6% (95% uncertainty intervals (CrI) 16.7 - 25.4), and cumulative mortality by 29.7% (95% CrI 25.8-33.8). A similar vaccine with ability to prevent symptoms (but not infection) will reduce peak incidence of symptomatic cases by 10.4% (95% CrI 8.4 – 13.0), and cumulative mortality by 32.9% (95% CrI 28.6 - 37.3). In the event of insufficient vaccine supply to cover all priority groups, model projections suggest that after keyworkers, vaccine strategy should prioritise all who are > 60, and subsequently individuals with comorbidities. In settings with weakest transmission, such as sparsely-populated rural areas, those with comorbidities should be prioritised after keyworkers.

**Conclusions:** An appropriately targeted vaccination strategy would witness substantial mitigation of impact of COVID-19 in a country like India with wide heterogenity. ‘Smart vaccination’, based on public health considerations, rather than mass vaccination, appears prudent.

**Strengths and limitation of this study**

- The model in this study is informed by age-dependent risk factors for SARS-CoV-2 infection among contacts, and is stratified by co-morbidities (diabetes and/or hypertension), and vaccination status.
- Data on mortality and large-scale contact tracing from within India, and the recent national sero-survey results were used, which constituted a major strength of this investigation.
- Distinguishing between ‘infection’ and ‘symptomatic disease ‘ preventing vaccines, the model was simulated under a range of scenarios for the basic reproduction number (R0).
- Should they have been available, real life country-specific data on excess risks of deaths due to comorbidities would have added strength to the presented model.
- Key priority group-specific data on social mixing and potential associated transmission was not available, and remained as a limitation.

## INTRODUCTION

COVID-19 has caused substantial morbidity and mortality worldwide, at levels not witnessed since the H1N1 influenza pandemic over a century ago.^1^ Non-pharmaceutical measures for its prevention such as hand hygiene, use of masks and maintaining physical distance during social interactions have played important roles in reducing the transmission of SARS-CoV-2, the causative agent. However, such measures, by themselves, are impractical for sustained suppression of viral transmission for long.^2–5^ In the meantime, development of vaccines against COVID-19 has progressed at an unprecedented pace. Promising results from phase 3 clinical trials of some of these candidates have emerged within a year from the publication of the whole genome sequence of SARS-CoV-2.^6^ Expectations on these vaccines range from prevention of infection and reduction of disease severity, to averting deaths among most at risk population groups.

Given that COVID-19 vaccines are already becoming available for distribution through public healthcare systems, many countries^7^ are now critically reviewing their vaccination plans. A major concern is how to effectively reach and engage a far larger number of individuals, the majority of whom are adults, than those typically covered under universal immunization programmes for children. Other important considerations include central storage facilities, the need for a cold chain to be maintained till vaccines are transported to the intermediary storage stations, and administered at the remotest vaccine session sites, and resource mobilization. Ethics and equity have also remained integral to these discourses^8^ where ‘vaccine nationalism’ has been examined in depth.^9^ The country of origin of a COVID-19 vaccine, production and procurement capacities of different countries, and concerns about inequitable global vaccine distribution; all compound such challenges.^9–11^

Against this background, and with a robust countrywide immunization program for children in place, India has come to the centre-stage of discussion related to COVID-19 vaccine. The second-most populous country in the world, India has accounted, at the time of writing, for 9% of COVID-19 cases reported worldwide, exceeded only by the United States and Brazil. Worth noting in this context is that India serves as a major source of vaccine production worldwide, accounting in 2019 for more than 60% of vaccines provided to low- and middle-income countries.^12^ In anticipation of mass vaccination against COVID-19, discussions were held on which population groups to be prioritised for vaccination. Three priority groups so far have been proposed based on public health considerations in India, (i) key workers, including healthcare professionals and other frontline workers, (ii) those over 60 years of age, and (iii) those aged between 24 to 60 years having comorbidities, as they are at increased risk of severe COVID-19 disease.^13^

In order to inform these discussions, we constructed a mechanistic mathematical model to estimate potential epidemiological impact of vaccinating the aforementioned priority groups, as well as to explore the effects of different strategies for vaccination, amongst these groups. The model is informed by age-dependent risk factors for SARS-CoV-2 infection among contacts. Mortality and contact data generated by a large-scale contact tracing study in India, ^14^ and the recent national sero-survey results^15^ have been used for this purpose. This modelling serves to illustrate some important considerations for vaccine planning, relevant to India as well as to other countries facing similar challenges.

## METHODS

India’s national serological survey completed its second round in August 2020, and estimated a seroprevalence of 7.1% (95% CI 6.2 – 8.2) at the country level, well under the theoretical herd immunity threshold for SARS-CoV-2. ^16^ The third round, completed in January 2021, estimated the seroprevalence to be 25%, underlining again the existence of a considerable proportion of vulnerable population in the country. Such findings suggested that a full easing of restrictions would lead to a rebound in transmission. (Indeed, several parts of the country are already seeing an increase in infections at the time of writing.) We modelled the potential impact of future vaccine rollout, in mitigating such a rebound. In particular, we examined which population groups should receive the vaccination first, under different scenarios for vaccine efficacy, and for the basic reproduction number, R0 (the latter, as estimated in the absence of any infection- or vaccine-induced immunity). We considered three different population groups for discussion as listed in figure 1, and in line with the ground reality in India.^17^ Consistent with ongoing practice, we assumed that key workers would receive vaccine first due to obvious ethical consideration (i.e. we excluded alternative scenarios where other groups might be prioritised over key workers). Holding this as a given, we examined the conditions under which those over 60 years of age should subsequently be prioritised over those with comorbidities, and vice versa.

**Figure 1.**
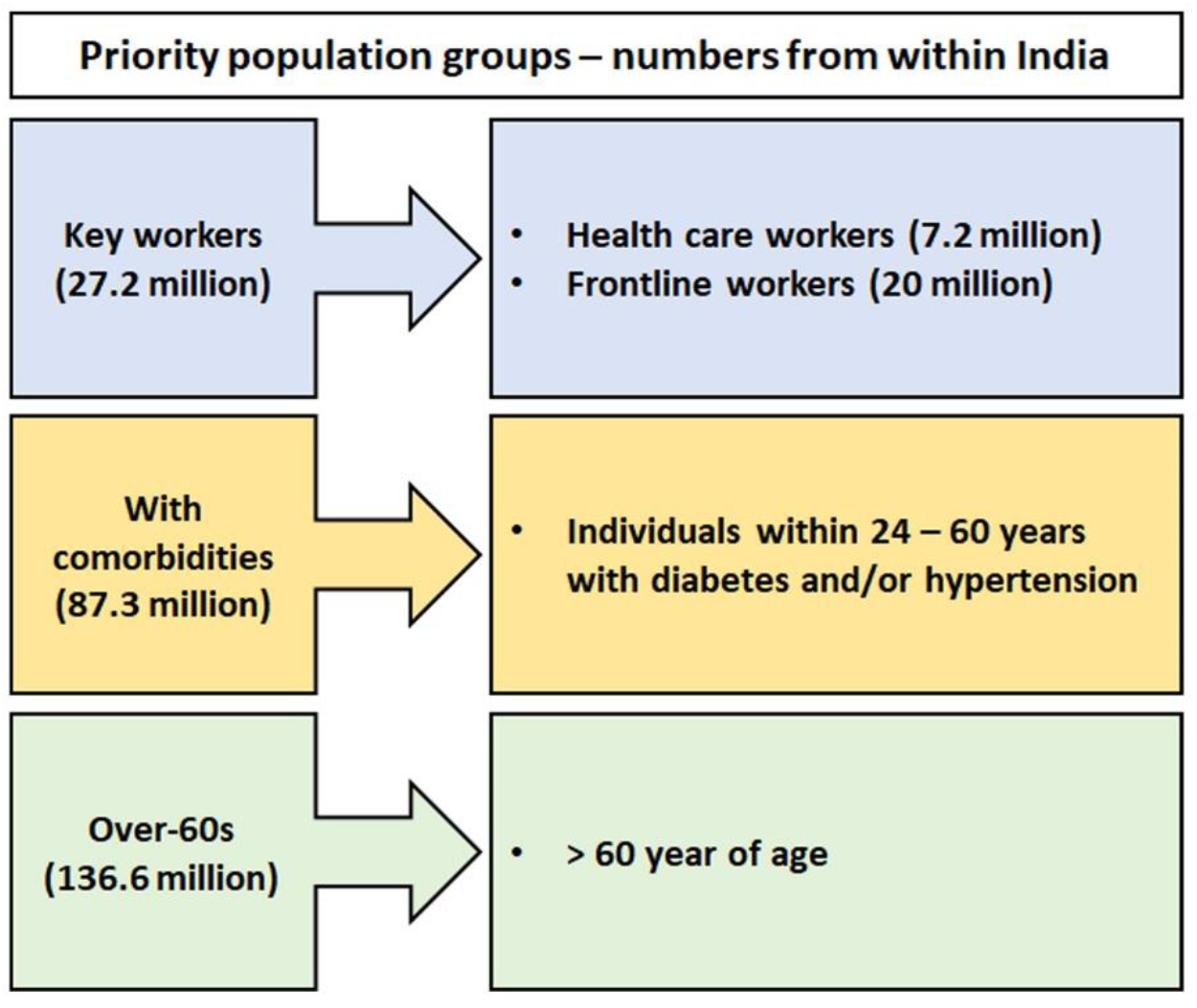
Priority groups of people in three different scenarios. Sources: healthcare workers (HCW)^30^, frontline workers (FW), those with diabetes and hypertension as co-morbidities^31^, those over 60 years of age^32^. As described in the main text, when modelling vaccination coverage amongst essential workers, our focus is on the epidemiological impact of doing so (we do not address, for the example, the potential impacts for healthcare continuity, of vaccination coverage in HCWs).

### Structure of the mathematical model

The model is a deterministic, compartmental framework, illustrated in figure 2 and shown in further detail in the supporting information. The model is stratified by different age groups (<24 year, 24 – 60 year, and >60 year); it is also stratified by comorbidities (diabetes and/or hypertension), and vaccination status. The model captures essential features in the natural history of SARS-CoV-2, including the role of asymptomatic infection, and the pronounced variations in disease severity, and mortality risk, by age (see table S1). To capture age-specific patterns of transmission (the ‘age-mixing’ matrix), we drew from recently published findings from a large contact tracing study in India.^14^ For the prevalence of comorbidities in different age groups, we drew the most recent estimates from the Global Burden of Disease study.^18^ As described below, we incorporated uncertainty in model parameters by defining plausible ranges for these parameters (see table S2), and then sampling from these ranges.

**Figure 2.**
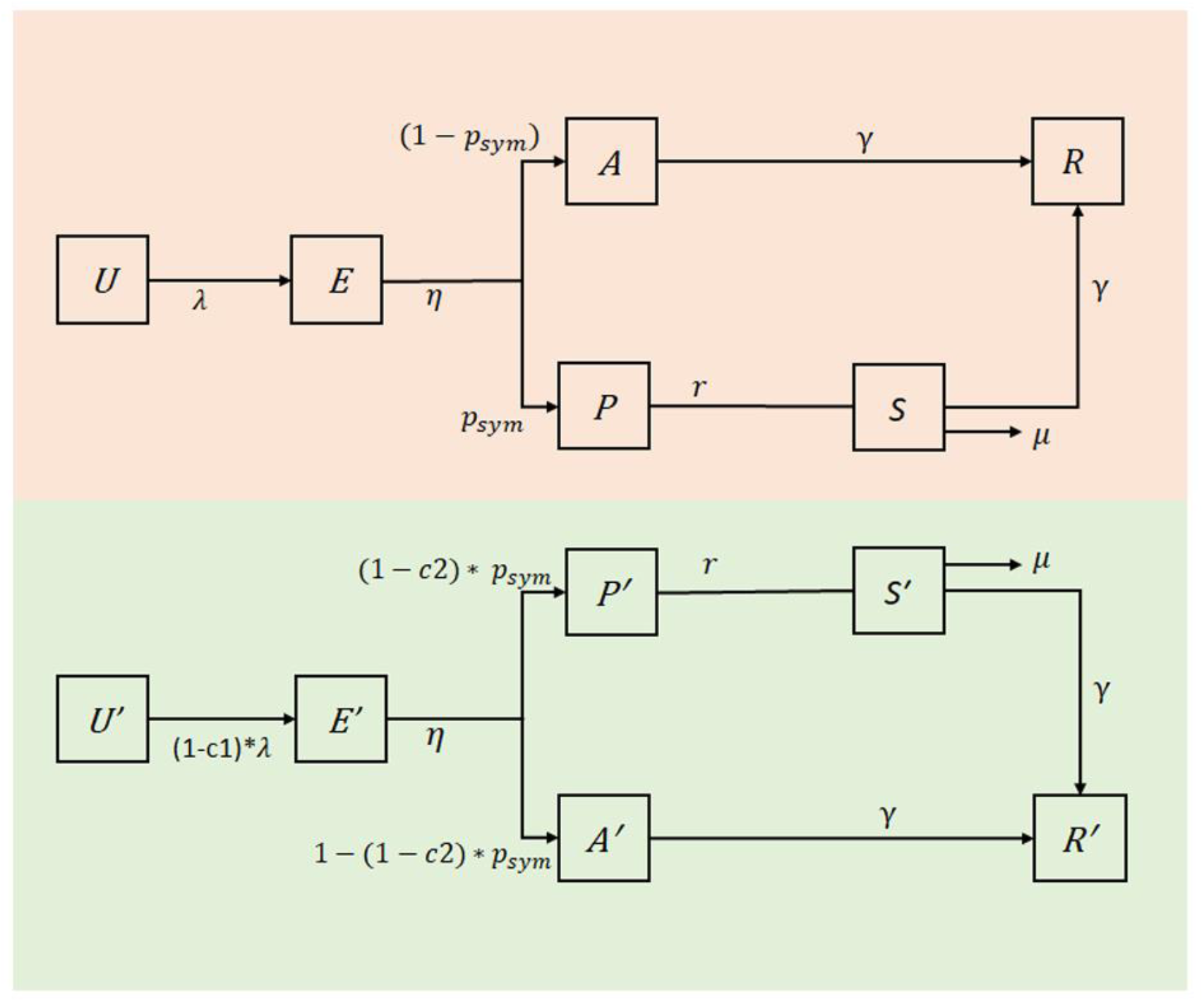
Illustration of the compartmental model structure. The top and bottom halves of the figure show unvaccinated and vaccinated subpopulations, respectively. Boxes represent compartments, and arrows represent flows between different stages of the clinical course of infection. Compartments are as follows: Uninfected (U); Exposed (E); asymptomatic but infectious (A); presymptomatic (P); symptomatic (S); recovered and immune (R). The terms *c*_1_, *c*_2_ represent effectiveness of, respectively, an infection preventing and symptomatic disease preventing vaccine. The term *μ* represents the per-capita hazard of mortality; see table S2 for a list of all other model parameters. This model structure is further stratified by age groups, and by presence/absence of comorbidities.

### Vaccination scenarios

We first modelled the potential impact of vaccination on incidence and mortality in all of the population groups identified in figure 1 (see table S3). Next, to examine prioritisation amongst these groups, we assumed that there is a sufficient vaccine stock to cover a given proportion *p* of the overall population. Assuming that key workers would receive first priority, we identified the second priority group in whom this amount of vaccine would lead to the greatest reduction in overall deaths, relative to a scenario of no vaccine; for any unused vaccine stock, we then identified how much of the remaining priority group would be covered with the remaining vaccine supply. We note that this analysis does not address temporal sequencing (i.e. which groups to vaccinate first in time). For instance, if model results suggest that the greatest mortality reductions could be achieved through vaccinating 100% of a given group and using remaining vaccine to immunise 25% of the remaining priority group, in practice the implementation of this coverage could proceed in both groups simultaneously. For simplicity in the modelling, for a given vaccine supply, we assumed that vaccination coverage is completed in advance of the epidemic (and can thus be modelled through initial conditions for the dynamical equations). We simulated deaths averted by vaccination, relative to a scenario of no vaccination. However, for comparison, we also modelled a ‘uniform’ strategy where vaccine supply is allocated proportionately amongst the two risk groups (those above 60 year of age and those between 24-60 year and with co-morbidity), rather than prioritising one over the other.

We repeated this analysis for a range of values for *p*, up to 18% of the population (the overall proportion of the population represented by the collective priority groups in figure 1). We also repeated this analysis for a range of values for R0 from 1.25 to 2.5, to capture the variability of transmission intensity across different settings within India, ranging from urban to rural.^15^

In addition, efficacy estimates for currently licensed vaccines – whether obtained through interim analyses or through bridging studies or trials in other countries - rely on symptomatic illness as an endpoint. The extent to which these vaccines may reduce infectiousness is currently unknown. In order to address these uncertainties, we modelled two types of vaccine: one that reduces susceptibility to infection with no effect on severity (an ‘infection-preventing’ vaccine), and one that reduces severity of infection (including mortality) with no effect on susceptibility (a ‘symptomatic disease preventing/modifying’ vaccine). In practice, it is likely that vaccines would have a combination of these two effects. By dichotomising their effects in this way, our analysis incorporates a range of possible scenarios for vaccine-induced protection.

Interim trial results from three separate vaccine candidates vary from 70% to 95%, ^19,20^ with other vaccine candidates also under consideration for use in India. As a conservative scenario for vaccine efficacy, given the complexity of implementation in a setting like India, we assumed a vaccine efficacy scenario of 60%. As a sensitivity analysis, we also simulated an alternative vaccine efficacy of 90% (Figs. S3 – S4). Regarding duration of vaccine-induced immunity, again conservatively a range from 3 months to 1 year was considered.^21^

### Uncertainty

For each model parameter relating to natural history of SARS-CoV-2 infection, we defined a plausible range of parameter values (see table S2). After drawing 5,000 independent samples from these ranges using latin hypercube sampling, we performed model projections on each sample and then estimated uncertainty on model projections, by designating the 2.5^th^ and 97.5^th^ percentiles as the 95% ‘uncertainty interval’ (CrI).

### Patient and public involvement

Patients and/or the public were not involved in the design, or conduct, or reporting, or plans of this research. However, dissemination plan of this investigation output will ensure availability of the results in the public domain and to inform public health discussions and debate.

## Results

Figure 3 shows illustrative model projections for the impact of vaccination to cover all of the priority groups listed in figure 1, in the example of the basic reproduction number R0 = 2. These results suggest that an infection-preventing vaccine with 60% efficacy could reduce peak symptomatic incidence by 20.6% (95% CrI 16.7 – 25.4) and cumulative mortality by 29.7% (95% CrI 25.8 – 33.8), relative to a scenario of no vaccination. A symptomatic disease preventing vaccine would have similar impacts on mortality, but little impact on symptomatic incidence. Results suggest that such a vaccine could reduce peak symptomatic incidence by 10.4% (95% CrI 8.4– 13.0) and cumulative mortality by 32.9% (95% CrI 28.6 – 37.3). Table 1 summarises these overall impacts, illustrating, for example, that vaccinating those over 60 year old would offer the greatest reductions in mortality per vaccinated individual, for both infection and symptomatic disease preventing vaccines.

**Table 1.** Summary of epidemiological impacts for the different scenarios shown in figure 3. Numbers show median estimates, while parentheses show 95% uncertainty intervals.

|  | Infection preventing vaccine |  |  | Symptomatic disease preventing vaccine |  |  |
| --- | --- | --- | --- | --- | --- | --- |
|  | Percentage reduction in peak symptomatic incidence | Percentage reduction in cumulative mortality | Number needed to vaccinate to avert one death | Percentage reduction in peak symptomatic incidence | Percentage reduction in cumulative mortality | Number needed to vaccinate to avert one death |
| (A) key workers (HCW + FW) | 4.8 (3.8 – 6.3) | 2.0 (1.4 – 2.8) | 1872 (1292 - 3031) | 2.3 (1.8- 3.1) | 2.0 (1.7 – 2.4) | 1877 (1226 - 3034) |
| (B) Key workers + Individuals with comorbidities (24 – 60 years) | 18.8 (14.9 – 23.6) | 11.8 (8.2– 15.7) | 320 (213 – 528) | 8.9 (7.1–11.9) | 13.6 (10.8 – 16.4) | 273 (179 - 460) |
| (C) Above two groups (A+B) + all individuals over 60 years of age | 20.6 (16.7 – 25.4) | 29.7 (25.8 – 33.8) | 127 (87 – 196) | 10.4 (8.4 – 13.0) | 32.9 (28.6 – 37.3) | 114 (76 - 184) |

**Figure 3.**
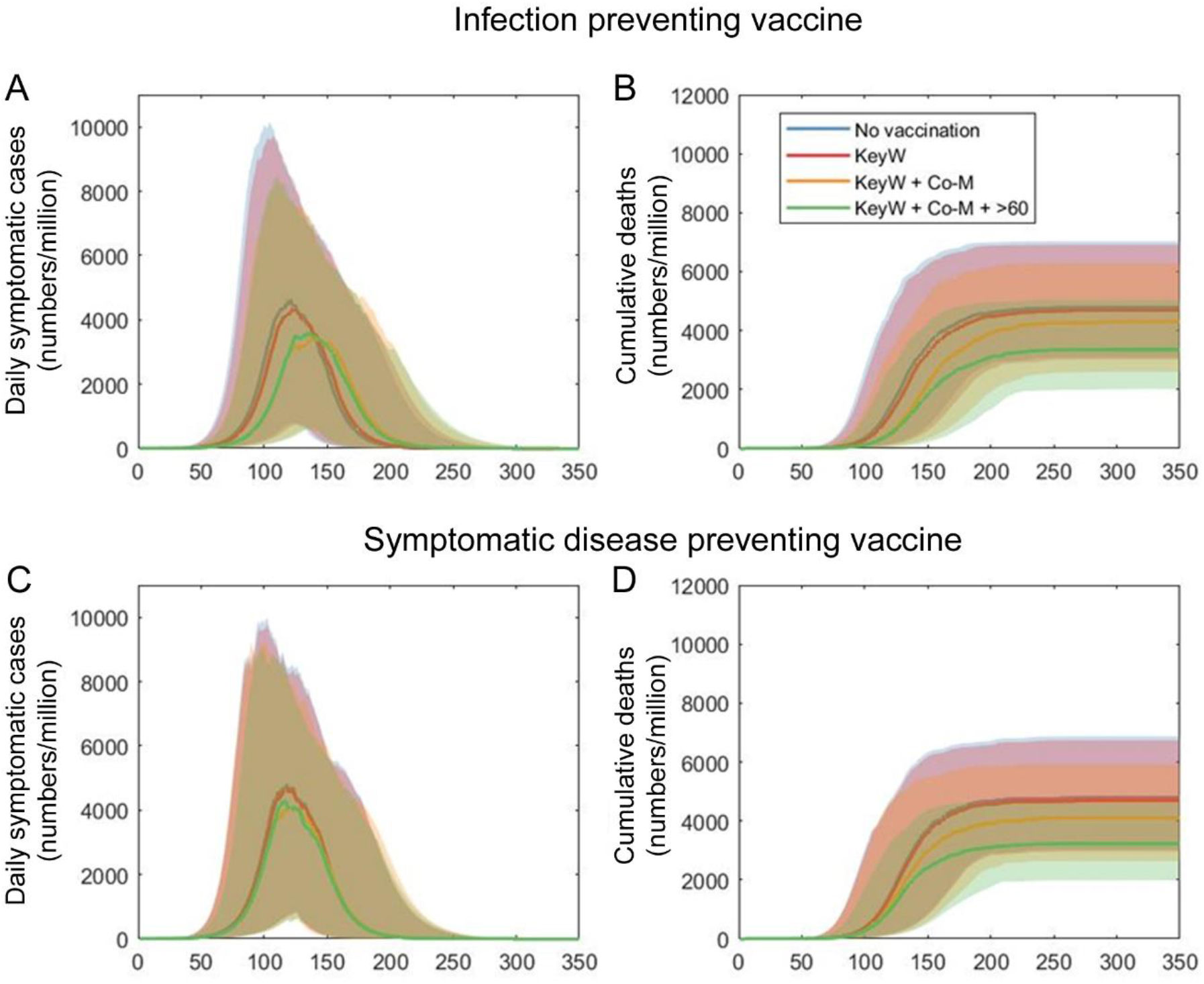
**Illustration of vaccine impact in each of the priority groups** listed in Figure 1. Scenarios are shown in the example of R0 = 2, assuming that vaccine coverage is completed at the same time as current restrictions being fully lifted. Upper row shows results for an infection-preventing vaccine, while the lower row shows results from a symptomatic disease preventing vaccine. Scenarios show vaccination coverage in different combinations of priority groups: Keyworkers (‘KeyW’); additionally including those with co-morbidities (‘Co-M’); and additionally including those over 60 years of age (‘>60’).All horizontal axes show days after vaccination and restrictions being lifted. Solid lines show central (median) estimates, while shaded areas show 95% uncertainty intervals as estimated by sampling uniformly from the ranges shown in table S2. The overall impact of vaccination in each of these scenarios is summarised in table 1, together with the amount of vaccines needed.

Even if there is ultimately sufficient vaccine production to cover all priority groups as shown in figure 1, in practice it is likely that supply would be staggered in the initial months of vaccine deployment, thus necessitating the identification of priority groups to target in these stages. Figure 4(A-C) shows illustrative results for an infection-preventing vaccine, for the optimal sequencing of priority groups. Most scenarios for R0, indicate prioritisation of those over 60 year old (those most at risk from severe outcomes of infection), before covering those with comorbidities (Figs. 4B,C). However, in settings with low transmission (R0 = 1.25), those with comorbidities should be prioritised over those older than 60 year (Fig. 4A). Figure 4(D-F) shows corresponding results for a symptomatic disease preventing vaccine; here again, the priority group after keyworkers is generally those over 60 year old (Figs. 4E,F) except in the low-R0 scenario (Fig. 4D), where those with comorbidities would instead be prioritised. In all cases, prioritising risk groups in this way would avert more deaths, or have comparable impact to, a ‘uniform’ strategy of allocating vaccines proportionally amongst risk groups (dotted grey line).

**Figure 4.**
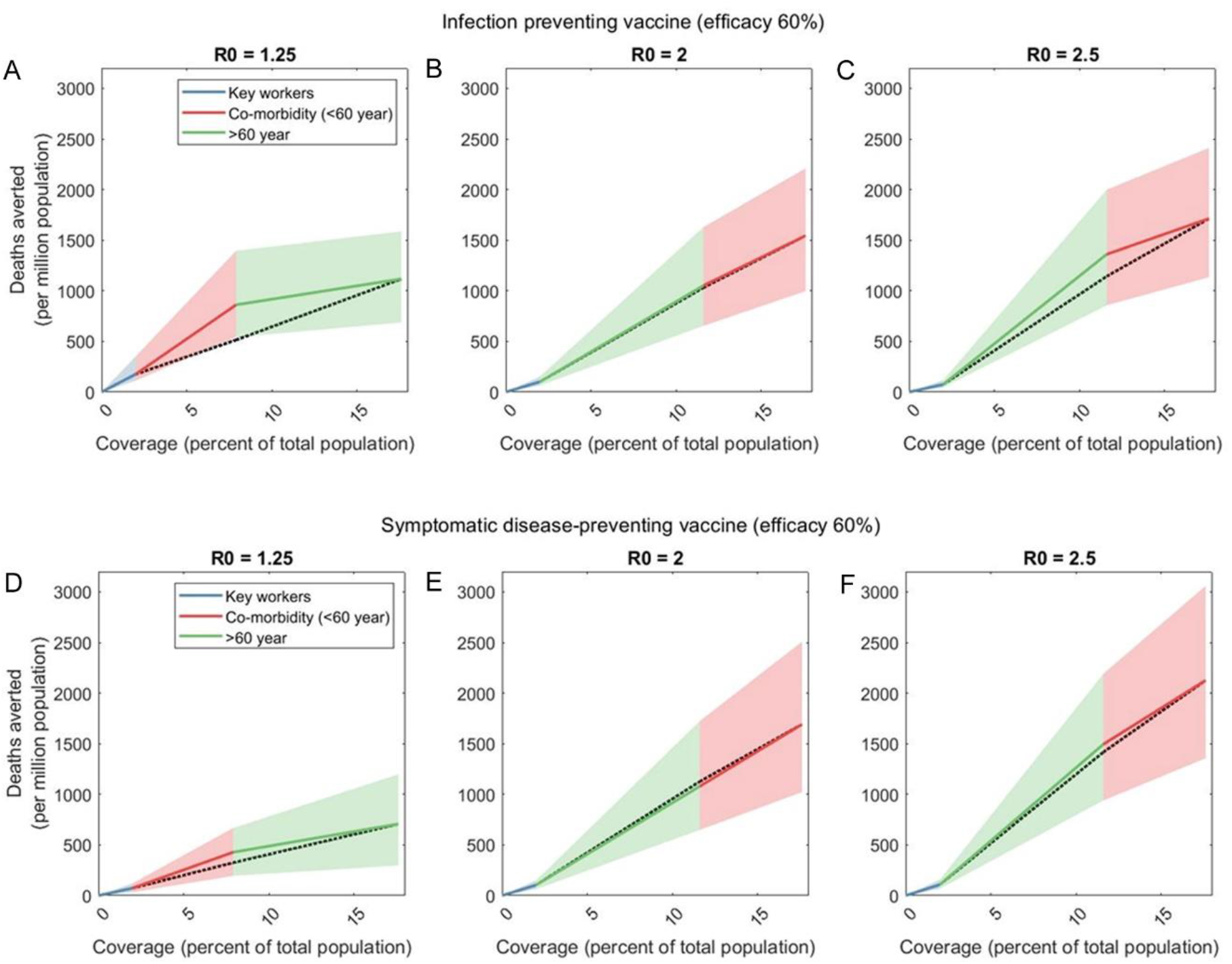
Optimal prioritisation strategies for an infection-preventing vaccine (A, B, C) and for a symptomatic disease preventing vaccine (D, E, F). For reference, dotted black lines in all plots show a ‘uniform’ strategy where available vaccines are allocated proportionately amongst the two risk groups, rather than prioritising one over the other (for clarity, uncertainty intervals not shown for this scenario). For the plots (A – C) we assume deployment of a vaccine having 60% efficacy in reducing susceptibility to infection, but no effect on development of symptoms following infection. Assuming keyworkers receive first priority, Figs.S1 – S2 in the supporting information show different strategies for subsequently prioritising those over 60 years old, vs those with comorbidities. Here, we show those strategies that are optimal for minimising the overall mortality, under different levels of vaccine coverage, and for different values of R0. For example,in the case R0 = 2, if initial vaccine supply is only enough to cover 10% of the population, then after covering keyworkers, these vaccines should be deployed preferentially amongst the over-60s (in green). If there is enough vaccine supply to cover 20% of the population, the optimal strategy would be to vaccinate the over-60s after keyworkers, and spending any remaining vaccine supply amongst those with comorbidities. Similar priorities apply for R0 = 2.5. However, for low-transmission settings (R0 = 1.25), those with comorbidities would be prioritised over the elderly. For the plots (D – F) we assume deployment of a vaccine having 60% efficacy in reducing symptoms and mortality following infection, but no preventive effect on acquiring infection. For such a vaccine, optimal prioritisation strategies are similar to those shown in plots (A-C).

## Discussion

Challenges that are particularly pressing in a country as large as India would persist even following the emergence of several vaccine candidates for COVID-19. The most contentions of them relate to rolling out of vaccines at population level. In this study, we have used a simple mathematical model of transmission dynamics, to show how vaccination efforts in the country might best be focused, in order to reduce mortality most effectively with a finite vaccine supply. Our results suggest that vaccinating all defined priority groups would have a substantial reduction in overall health burden, compared to a scenario of no vaccination, and complete lifting of restrictions. Such a strategy could reduce peak symptomatic incidence by about 21%, and cumulative mortality by about 30% .

In terms of prioritisation of population groups, our results show how the most efficient use of a given vaccine supply is shaped by transmission intensity (R0), whether for infection- or symptomatic-disease-preventing effects of the vaccine (figures 4). Conceptually, the fundamental dynamics underpinning these results arise from interactions between ‘direct’ effects of immunisation (i.e. the protection amongst those receiving the vaccine) and ‘indirect’ effects (i.e. the population-level benefits of general reductions in transmission). While in practice any vaccine is likely to exert a combination of both the effects, our work highlights that, for a vaccine supply sufficient to cover 18% of the population, direct effects would generally take precedence over indirect effects, in deciding prioritisation. Thus vaccination rollout should generally prioritise those most at risk of severe outcomes of infection; in the present case, the elderly. However, only in the lowest-transmission settings would those with comorbidities be prioritised over the elderly. As those with comorbidities include young adults, who have greater contact rates than the elderly, vaccinating this group would raise stronger indirect effects; it is in low-R0 scenarios that such effects would be as important as direct effects.

Our results highlight the need for further data to help inform strategic priorities, both on transmission in real world settings (i.e. R0 in any given setting) and vaccine effect on transmission. On the first of these, although clinical trials so far have focused on symptomatic illness as an endpoint, interim findings for at least one vaccine candidate suggest the potential for reduced transmission as well.^19^ However, further data are needed, for example through trial designs following up household cohorts to assess the risk of transmission amongst close contacts, and how this risk is affected by vaccination.

Alternatively, a better understanding of how viral load correlates with SARS-CoV-2 transmission could allow better interpretation of available trial results, in terms of transmission risk. ^22,23^ On the latter point mentioned above, mathematical and statistical models – similar to those we have presented here - have been used to estimate R0 for SARS-CoV-2 in different settings, and may also be informative in the Indian context.^14^ We note that in a country as large and complex as India, there will be a need for locally-tailored, locally-relevant estimates. As an indication of varying transmission intensity across the country, the second national serosurvey reported 16% seroprevalence of SARS-CoV-2 antibody among those living in urban slums; 8% among those living in urban non-slum setting; and 4% in rural settings.^15^ Such variation is likely to be driven by factors such as population density, and indeed may call for different prioritisation strategies in different settings. For example, scenarios of R0 = 1.25 and 2.5 may be appropriate, respectively, in rural and urban slum settings. In all of these considerations, robust surveillance data – including at the level of hospitalisations and mortality – would be invaluable in refining model estimates.

As described above, our analysis does not explicitly address temporal sequencing, i.e. which groups to cover first: for simplicity, we modelled vaccination coverage as being completed in advance of the epidemic, concentrating on identifying the groups who would have the most impact on mortality if receiving the vaccine. Nonetheless, our results can be interpreted in terms of temporal sequencing as well; in particular, even in the scenario where there is sufficient vaccine to cover 100% of the identified risk groups, critical challenges of prioritisation will arise in the event that an epidemic begins during the course of vaccine rollout. In such an event, the ‘effective’ coverage is simply the number of individuals who have been successfully immunised before being exposed to infection. Framed in this way, our results can therefore also be interpreted as the sequence of prioritisation that should be implemented, in order to maximise vaccine impact under a given amount of effective coverage.

As with any modelling study, our analysis has limitations to note, which should be regarded as illustrating the importance of different factors for policy decisions, and not as a predictive framework. It is subject to various uncertainties, for example, the increased risk of death as a result of comorbidities. Further data on these excess risks will be valuable in refining our findings. In considering the key worker population, although we incorporated vaccination coverages consistent with the size of this population, we did not explicitly capture the broader societal impact of failing to vaccinate these individuals, another important area for future work. Finally, an important uncertainty relevant to our current work is the dynamics of immunity, whether induced by vaccination or by infection. For example, there is evidence that memory B-cells and neutralising antibodies persist at detectable levels in blood for months post-infection ^24–26^. Despite important recent advances in understanding implications for disease outcome upon reinfection ^27^, there remains much uncertainty, including on the role of the cellular immune response ^28^. A recent modelling study showed how immune mechanisms could mediate a decline in the severity of COVID-19 as it becomes endemic in the coming years ^29^, but it remains unclear how current licensed vaccines, in India and elsewhere, might shape these dynamics. Addressing these issues are beyond the scope of our current work, which focuses on the implications of vaccination for immediate mitigation of health burden: nonetheless, these again represent important areas for future work to address.

In conclusion, models such as the one presented in this article can generate useful program insights. In practice the gains, as projected by the model due to vaccination of select population groups in real life settings, would enhance from other prevention measures at the population level such as use of masks and maintenance of physical distance during social interactions. Such a synergy is expected to yield further dampening of SARS-CoV-2 transmission. We therefore conclude that rational and focused vaccination approaches, as outlined in this article, in the context of Indian COVID-19 epidemic makes for a smarter public health choice than mass vaccination.

## Data Availability

The model code and dataset are publicly available at https://github.com/sandipccmb/COVID-19-vaccination-strategy.

## Author contributions

SP and BB conceptualised the study; SM, NA and SP developed the modelling approach and SM performed the modelling. All authors analysed and interpreted the results; SM and SP wrote a first draft of the manuscript, and all authors contributed to the final draft and approved the version for submission to the journal.

## Funding

Authors (SM, BB and SP) acknowledge funding from the Indian Council of Medical Research, and NA acknowledges funding from the UK Medical Research council. No additional funding or grant support was utilised for execution of this study by the authors who remained supported by their respective institutes of affiliation as indicated while independently carrying out the present study. The respective institutions of the authors had no financial interest in the investigational work.

## Competing interests

The authors declare no competing interests.

## Ethical approval

Not required.

## Notes

### Competing Interest Statement

The authors have declared no competing interest.

