## Supplementary materials for "INDIA’S PRAGMATIC VACCINATION STRATEGY AGAINST COVID-19: A MATHEMATICAL MODELLING BASED ANALYSIS"

### 1. Model specification

We developed a deterministic, compartmental model of SARS-CoV-2 transmission and disease course with three different age groups: <24 year, 24 - 60 year and >60 year, and further stratified by the presence of comorbidities. In all equations that follow, state variables (e.g. $U$, $E$ etc) denote the respective *proportions* of the total population in the corresponding states. Thus at time zero (prior to the epidemic), all state variables sum to 1. In this way, the model results can be applied to different administrative scales within India (e.g. districts), regardless of the actual population size involved. Accordingly, all model results are shown as population rates, e.g. deaths per million population (Figure 3, main text).

*Governing equations*

Model compartments are listed in Table S1, and model parameters listed in Table S2. Governing equations for the non-vaccinated population are as follows, where subscript $i$ denotes age group, and subscript $j$ denotes comorbidity group:

Uninfected ($U$):

$$\frac{dU_{ij}}{dt} = -\lambda_i U_{ij}$$

Exposed but not yet infectious ($E$):

$$\frac{dE_{ij}}{dt} = \lambda_i U_{ij} - \eta E_{ij}$$

Asymptomatic and infectious ($A$):

$$\frac{dA_{ij}}{dt} = \eta \left(1 - p^{(sym)}\right)E_{ij} - \gamma A_{ij}$$

Presymptomatic and infectious ($P$):

$$\frac{dP_{ij}}{dt} = \eta \, p^{(sym)} \, E_{ij} - r \, P_{ij}$$

Symptomatic and infectious ($S$):

$$\frac{dS_{ij}}{dt} = rP_{ij} - \mu_{ij} \, S_{ij}$$

Recovered and partially immune ($R$):

$$\frac{dR_{ij}}{dt} = \gamma(A_{ij} + S_{ij})$$

A key parameter here is $p^{(sym)}$, the proportion of infected individuals developing symptoms.

Corresponding equations apply for the vaccinated compartments, but with primes distinguishing these compartments (e.g. $U'$). Additionally for this population, the term $p^{(sym)}$ is replaced by $(1 - c_2)p^{(sym)}$, where $c_2$ is vaccine efficacy in preventing disease.

For the force-of-infection experienced by non-vaccinated individuals, we have:

$$\lambda_i = \sum_{k,l} \beta\, m_{ik} \{[S_{kl} + k\,(A_{kl} + P_{kl})] + [S'_{kl} + k\,(A'_{kl} + P'_{kl})]\}$$

And for vaccinated individuals:

$$\lambda'_i = (1 - c_1)\, \lambda_i,$$

where $c_1$ is the effect of the vaccine on reducing susceptibility to infection.

Overall, the value of the basic reproduction number ($R_0$) for this model is proportional to the value of $\beta$, the rate-of-infection attributable to symptomatic individuals (noting that $k$ acts as an adjustment for a/pre-symptomatic individuals). As described below, we controlled for $R_0$ by adjusting the value of $\beta$ accordingly.

| State symbol | Meaning |
|---|---|
| $U_i$ | Uninfected (i = 1, 2, 3 indicating three age groups) |
| $E_i$ | Exposed |
| $A_i$ | Asymptomatic |
| $P_i$ | Pre-symptomatic |
| $S_i$ | Severe symptomatic |
| $R_i$ | Recovered |

**Table S1 List of state variables**

<table>
<tr><th>Parameter</th><th>Meaning</th><th colspan="3">Values</th><th>Source/Remarks</th></tr>
<tr><td>$\beta$</td><td>Transmission rate</td><td colspan="3">0.079 – 0.16</td><td>Calculated using next-generation matrix as described in ref[1]. Value shown here is to yield R0 = 1.25 – 2.5.</td></tr>
<tr><td>$\eta$</td><td>Amongst those exposed, rate of developing infectiousness</td><td colspan="3">(1/3 – 1/5) /day</td><td>Corresponds to an average latent period of 3-5 days: together with the period of presymptomatic transmission (see $r$ below), corresponds to an overall average incubation period of 4-6 days [2]</td></tr>
<tr><td>$p^{(sym)}$</td><td>Proportion developing symptoms</td><td colspan="3">1/3 – 2/3</td><td rowspan="2">Wide variation noted in individual studies and meta-analysis [3–5]</td></tr>
<tr><td>$k$</td><td>Relative infectiousness of asymptomatic vs symptomatic infection</td><td colspan="3">2/3 – 1</td></tr>
<tr><td>$r$</td><td>Rate of developing symptoms</td><td colspan="3">1 /day</td><td>Assumption, corresponds to mean pre-symptomatic duration of 1 day</td></tr>
<tr><td>$\gamma$</td><td>Recovery rate</td><td colspan="3">0.2 /day</td><td>Assumption, corresponds to mean infectious period of 5 days[6]</td></tr>
<tr><td>$f$</td><td>Fold-increase in case fatality rate as a result of comorbidities (diabetes and/or hypertension)</td><td colspan="3">2.5</td><td>Drawn from recent systematic review [8]</td></tr>
<tr><td></td><td>Age groups</td><td><24 year</td><td>24-60 year</td><td>>60 year</td><td></td></tr>
</table>

| | | | | | |
|---|---|---|---|---|---|
| $CFR_i$ | Case fatality rate in age group $i$ in absence of comorbidities | 0.1% | 1.45% | 10.9 % | Drawn from a recent study from two Indian States.[9] |
| $\mu_i$ | Mortality rate for severe cases | 0.0002 /day | 0.0029 /day | 0.02 45 /day | Hazard rates of $\mu_i$ are calculated to yield case fatality rates, using: $CFR_i = \mu_i/(\mu_i + \gamma)$. Uncertainty in the mortality hazards are considered +/-25%. |
| $N_i$ | Population (India) | 634 mn | 614 mn | 131 mn | Extrapolated from the Census of India 2011[10] |
| $m_{ij}$ | Connectivity matrix between age group i with age group j | 1.37<br>2.52<br>0.28 | 1.43<br>2.90<br>0.34 | 0.05<br>0.01<br>0.02 | Drawn from ref.[9] Uncertainty in the each element of the contact matrix is considered +/-25%. |

**Table S2: Parameters used in the model simulation**. There remains much uncertainty about parameters relating to SARS-CoV-2 natural history, e.g. infectiousness of asymptomatic people relative to symptomatic ones and, duration of pre-symptomatic period etc. In this study we adopted a range of parameter values to reflect this uncertainty in our model projections (figure 3-5, main text).

### 2. Model execution

Using latin hypercube sampling, we drew 5,000 independent samples from the parameter ranges listed in Table S2. For each sample, and under given scenarios for $R_0$ and vaccine coverage, we then performed the following steps:

1. Control for the basic reproduction number ($R_0$), as follows:
    a. In the absence of any vaccination coverage or prior immunity, use analytical methods described in (ref[1]) to calculate the value $\rho$ of the reproduction number when $\beta = 1$.
    b. Set $\beta = R_0/\rho$, thus yielding the scenario-specified value of $R_0$ for the basic reproduction number.
2. Construct initial conditions for the dynamical system, as follows:
    a. Construct a disease-free population with no prior immunity except for those who have been vaccinated (the latter, in line with the specified scenario for vaccination coverage).
    b. Introduce infection by displacing 1 individual from the susceptible, unvaccinated adult population, to the symptomatic, unvaccinated adult compartment (the specific choice of characteristics for this seeding infection are not important for the model outcomes we analyse).
3. Simulate the system of equations listed in section 1, until there are no further new infections.
4. Record the cumulative deaths that occurred over the simulation period.

We repeated these steps for each of the 5,000 samples, to obtain a corresponding number of estimates for cumulative deaths. We then estimated uncertainty by taking 2.5$^{th}$, 50$^{th}$ and 97.5$^{th}$ percentiles over these samples.

### 3. Priority population groups for vaccination – further information

| Category | Numbers | | Source |
|---|---|---|---|
| **Number of healthcare workers (HCW)** | | | |
| HCWs (qualified) | 3827820 | | Karan et al (2019) [11] |
| Support workers | 1245878 | | |
| HCW (without requisite qualifications) | 2084185 | | |
| Total | 7157883 | | |
| **Frontline workers (FW)** | | | |
| | **Active** | **Reserve** | |
| Armed forces | 1443921 | 1155000 | Information available in public domain [12,13] |
| Paramilitary forces | 87000 | | |
| Central Armed Forces and Others | 1403700 | 987800 | |
| Municipal workers | 15000000 | | |
| Total | 20077421 | | |
| **Co-morbidity (diabetes and/or hypertension)** | | | |
| Population < 24 year of age with at-least one comorbidity | 17801137 (2.8% population in this age group ) | | WHO SAGE report, 2013[14] |
| Population 24 – 60 year of age with at-least one comorbidity | 87283375 (14.3% population in this age group ) | | |
| Population >60 year of age with at-least one comorbidity | 58726385 (43.0% population in this age group ) | | |
| **Elderly population** | | | |
| Population > 60 year of age | 136620434 | | Extrapolated from the Census of India 2011[10] |

**Table S3: Priority population groups for vaccination.**

### 4. Additional model outputs

Figure 4 in the main text shows model results for how priority groups might be sequenced, to gain maximum impact (lives saved) from a limited vaccine supply. While the figure shows only the 'optimal' scenario, Figures S1 below shows all 2 possible scenarios for the order in which vaccination is deployed amongst the priority groups, in the case of an infection-preventing vaccine, and assuming that keyworkers receive first priority. Of these, the optimally efficient scenario is selected as that with the greatest gradient (lives saved per person vaccinated) at each stage, i.e. the scenario having the most concave shape. Figures S2 show corresponding results in the case of a disease-preventing vaccine.

Scenario definitions are as follows:

**Scenario 1:** Key workers → Co-morbidity → Elderly

**Scenario 2:** Key workers → Elderly → Co-morbidity

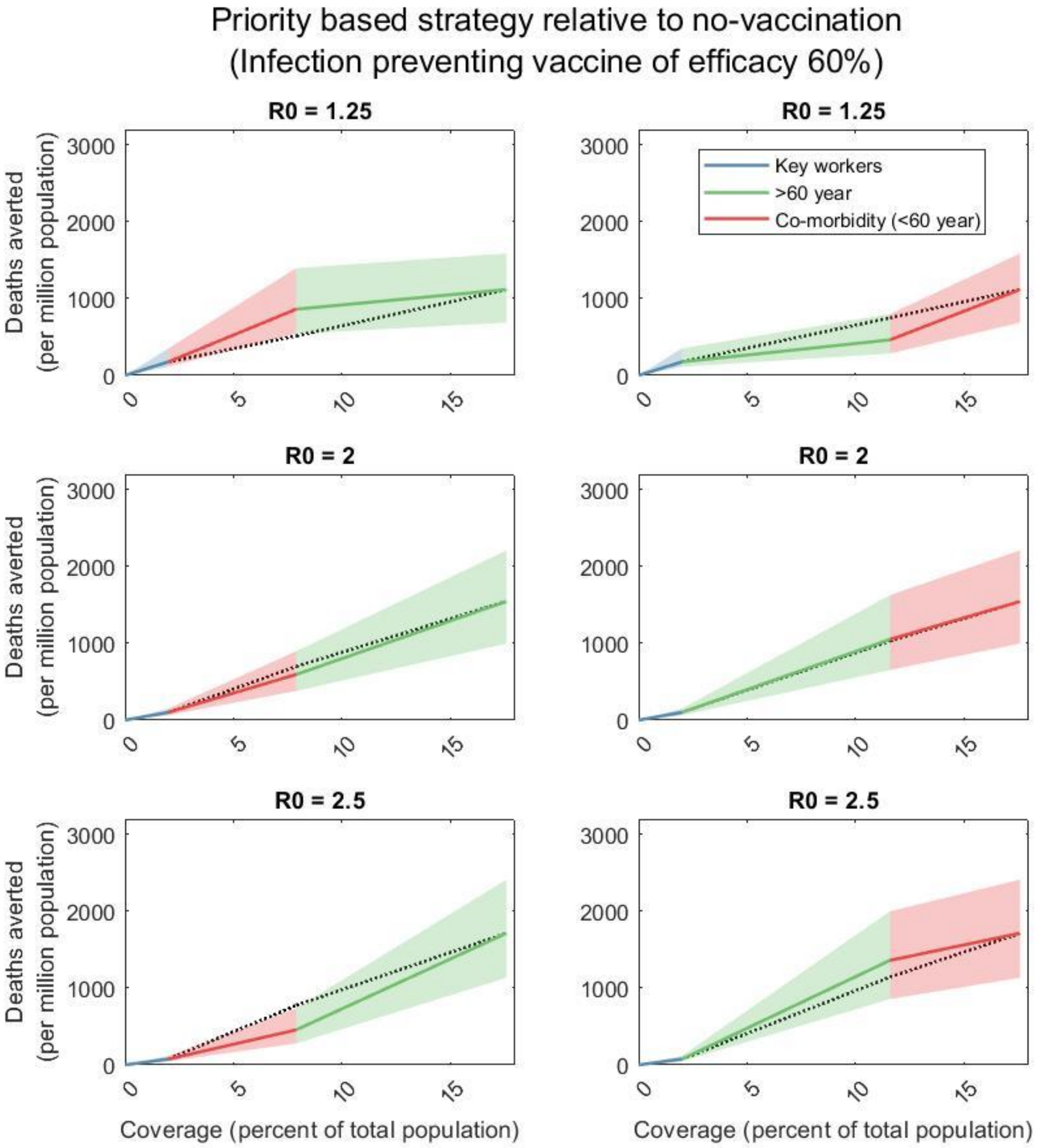

**Figure S1**. Scenarios for the order in which vaccination is deployed amongst the priority groups, in the case of an infection-preventing vaccine of efficacy 60%. We assume that keyworkers always receive first priority, and present scenarios for the prioritisation of the remaining two groups. As in the main text, dotted black lines show a ‘uniform’ strat R0 = 2.5 where available vaccines are allocated proportionately amongst the two risk groups, rather than prioritising one over the other.

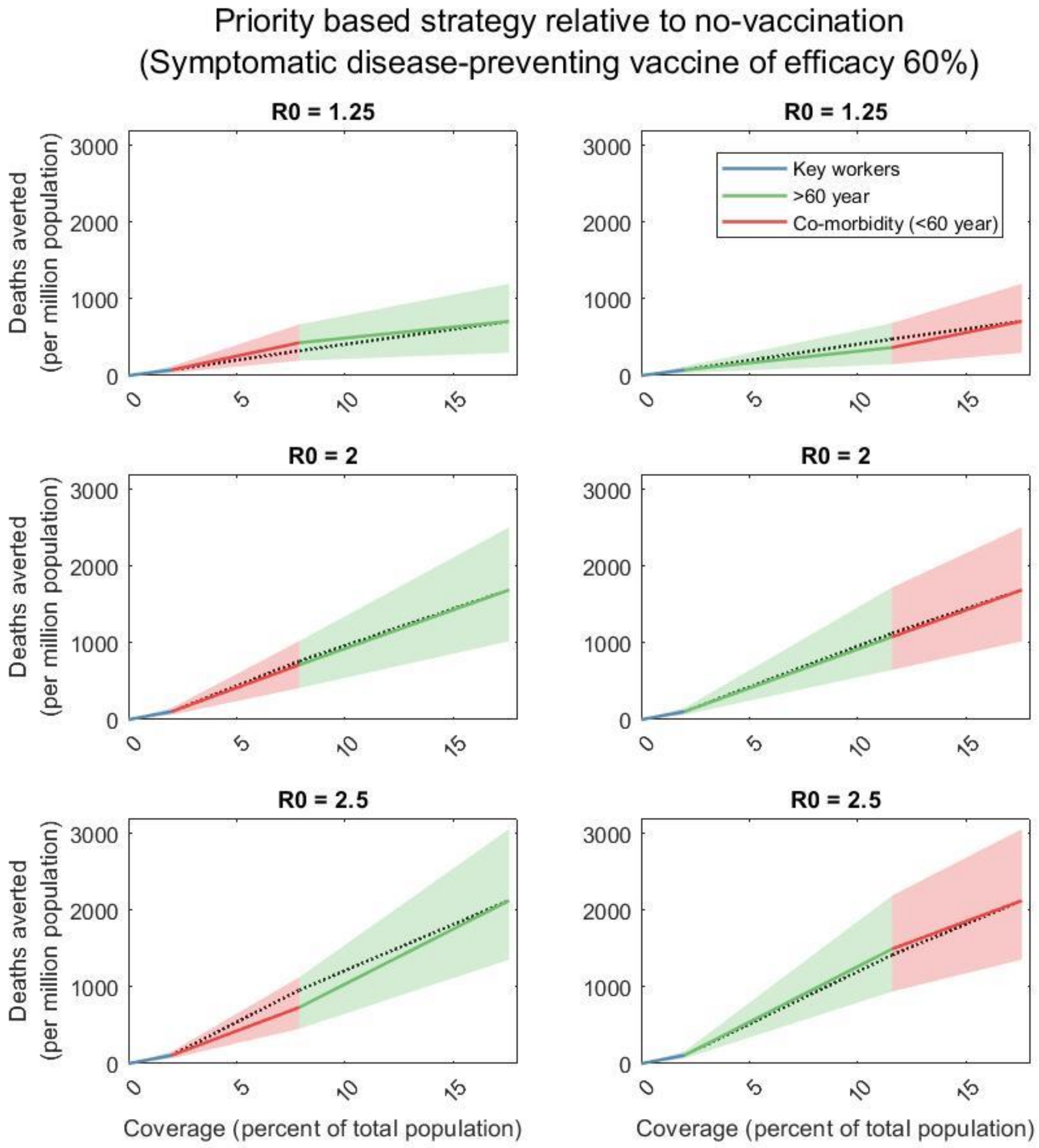

**Figure S2**. Scenarios as in figure S1, in the case of a disease-preventing vaccine of efficacy 60%.

### 5. Sensitivity analysis to vaccine efficacy

While results in the main text assumed (conservatively) a vaccine efficacy of 60%, below we present alternative results for 90%, showing that Figures 4 and 5 in the main text remain qualitatively unchanged.

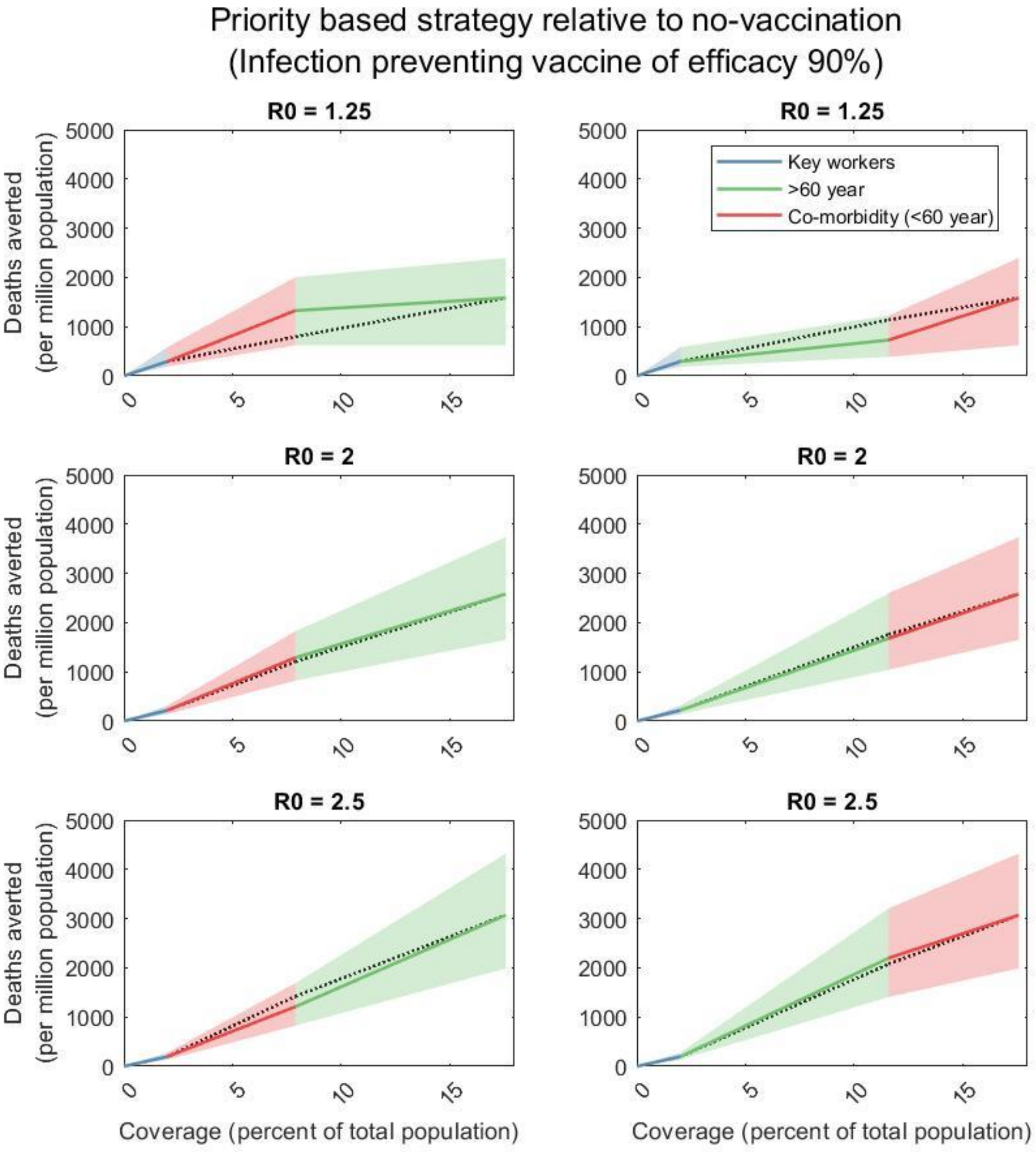

**Figure S3**. Scenarios for the order in which vaccination is deployed amongst the three priority groups, in the case of an infection-preventing vaccine of efficacy 90%.

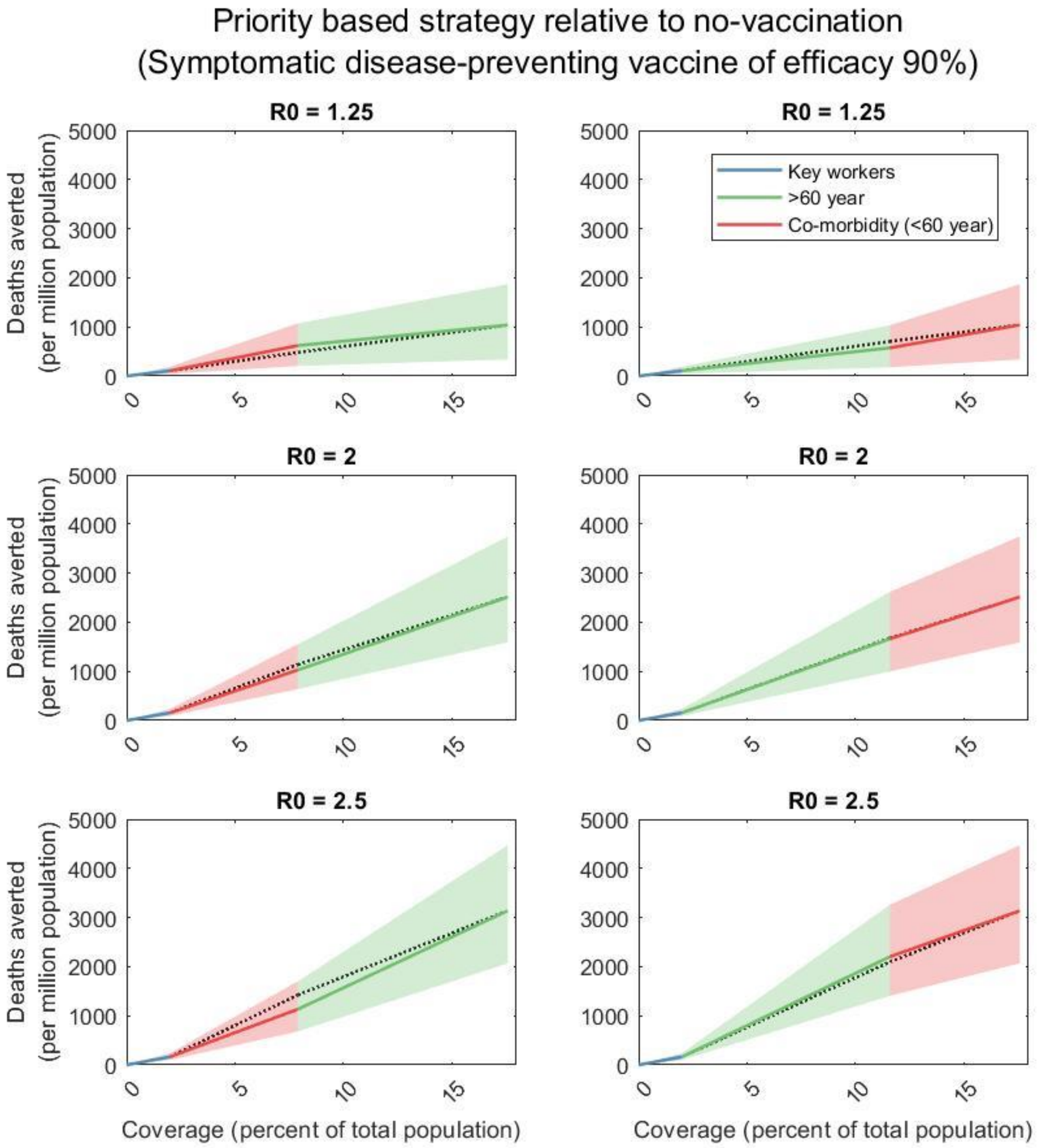

**Figure S4**. Scenarios as in figure S3, in the case of a disease-preventing vaccine of efficacy 90%.
